# Exercise Crystal: simulations that drive National IHR Focal Point capacity strengthening

**DOI:** 10.1101/2025.02.07.25321836

**Authors:** Laura Goddard, Qiu Yi Khut, Gina Samaan

## Abstract

The International Health Regulations (IHR 2005) require States Parties to designate a National IHR Focal Point (NFP) for timely IHR communications with the World Health Organization (WHO) and, following recent amendments, to designate a National IHR Authority to coordinate IHR implementation within States Parties. Since 2008, the WHO Regional Office for the Western Pacific has been supporting the strengthening and testing of NFP functionality through an annual simulation exercise known as IHR Exercise Crystal. This study analyses NFP performance during IHR Exercise Crystal over a 16-year period to inform Member States’ planning for NFP capacity strengthening, in the context of the recent IHR amendments. Exercise monitoring data were analysed using descriptive statistics across six key NFP performance indicators. Findings show consistently high NFP accessibility via email (mean: 99%) but suboptimal NFP accessibility via telephone (mean: 64%). NFP participation in tele- and videoconferences improved over time (mean: 73%), as did the proportion of NFPs notifying WHO of simulated events (mean: 80%) and contributing information to Event Information Site postings (mean: 77%). Multisectoral communication remained variable with no clear trend (mean: 73%). This demonstrates that significant progress has been made and that opportunities remain to further enhance NFP functionality, particularly in the areas of NFP accessibility and multisectoral coordination. It is critical that States Parties continue strengthening and testing NFP functionality through simulation exercises and other capacity-building to ensure effective IHR implementation. Furthermore, States Parties should develop, test and maintain up-to-date SOPs to support the clear demarcation of roles and responsibilities between the NFP and the National IHR Authority.

## INTRODUCTION

The International Health Regulations (IHR 2005) defines countries’ rights and obligations in handling public health events and emergencies that have the potential to cross borders.^1^ Signatory States Parties commit to developing core capacities for public health emergencies and, under Article 4, are required to designate or establish a National IHR Focal Point (NFP) for communications with the World Health Organization (WHO). The NFP must be accessible at all times for communications with WHO IHR Contact Points and is responsible for sending information to WHO on behalf of States Parties, as well as disseminating information to, and consolidating input from, relevant sectors of the administration of the State Party concerned.^1,2^ In the case of territories and areas, States Parties may establish IHR Contact Points specific to each territory or area for the purpose of IHR communications, though this is not mandatory under IHR (2005).

IHR communications and the role of the NFP are central to the IHR and to global health security. In the recent amendments to the IHR, the requirement to designate or establish an NFP has remained unchanged. However, States Parties must now designate or establish a National IHR Authority, which may be the same entity as an NFP or a different entity, to coordinate implementation of the Regulations within the State Party.^3^ The designation of a National IHR Authority may therefore impact on current NFP practices or operations in some countries.

In the WHO Western Pacific Region, IHR implementation is supported through *the Asia Pacific Health Security Action Framework* (APHSAF).^4^ Progress in IHR implementation, including NFP and other IHR core capacities, is assessed and monitored through the mandatory States Parties Self-Assessment Annual Reporting Tool (SPAR) and voluntary assessments, including Joint External Evaluations (JEE), simulation exercises and intra/after action reviews.^1,5^ These assessments help to identify strengths and areas for improvement and then translate them into priority actions as part of national planning to build country capacities.^6^

In the Western Pacific Region, the functioning of NFP capacities is routinely assessed through an annual simulation exercise organized by the WHO Regional Office for the Western Pacific (Regional Office). IHR Exercise Crystal has been running since 2008 and aims to test IHR communication channels and familiarize NFPs and WHO staff with the IHR communication system.^7^ Findings from the exercise can inform actions needed to strengthen NFP functionality and should be triangulated with other assessments to better understand the capacity level of countries and to implement priority actions at national and subnational levels.^5^

This analysis describes the performance of IHR NFPs and Contact Points in the Western Pacific Region during IHR Exercise Crystal over time, with the aim of informing Member States’ planning as they prepare for implementing the IHR amendments.

## METHODS

### Study design

We conducted a descriptive analysis of IHR NFP and Contact Point performance during IHR Exercise Crystal over a 16-year period (2008–2024, excluding 2009).

### Study population and setting

The Regional Office works with health authorities from 37 countries and areas, of which 22 are Pacific island countries and areas, totalling more than one quarter of the world’s population.^8^ The region is very diverse; significant variation exists in Member States’ geography, demographics, health systems and services, disease burden and disaster risk profiles.^9–11^ There are, however, many common challenges and approaches that countries and areas in the region share, including a common approach to IHR implementation and capacity development through APHSAF.^4^

### Data sources and analysis

Data on NFP performance during IHR Exercise Crystal were extracted from exercise reports and monitoring datasets for the years 2008–2024. Monitoring and evaluation data have been collected each year of IHR Exercise Crystal to measure IHR NFP and Contact Point performance against the exercise objectives (Table 1); however, where monitoring datasets were not available (years 2008, 2010, 2019 and 2021), only exercise reports were used.

**Table 1.** IHR Exercise Crystal objectives by year, 2008–2024.

| Objectives | 2008 | 2009 | 2010 | 2011 | 2012 | 2013 | 2014 | 2015 | 2016 | 2017 | 2018 | 2019 | 2020 | 2021 | 2022 | 2023 | 2024 |
| --- | --- | --- | --- | --- | --- | --- | --- | --- | --- | --- | --- | --- | --- | --- | --- | --- | --- |
| Validate the accessibility of NFPs using registered contact details | X | N/A | X | X | X | X | X | X | X | X | X | X | X | X | X | X | X |
| Practice and test the IHR notification process | – | N/A | X | X | X | X | X | X | X | X | X | X | X | X | X | X | X |
| Cross-sectoral communication between NFPs and national counterparts | – | N/A | – | – | – | – | X<sup>a</sup> | – | X<sup>b</sup> | X<sup>c</sup> | X | X | X | X | X | X | X |
| Improve NFP understanding of the IHR communication system | – | N/A | – | – | – | – | – | X | X | X | X | X | X | X | X | X | X |
| Test the use of tele- or videoconferencing | – | N/A | X<sup>d</sup> | X | X | – | – | X<sup>e</sup> | X<sup>d</sup> | X | X | X | X | X | X | X | X |
| Additional objectives | 1, 2 |  | 1 |  |  | 3 |  | 4 | 5, 6 | 4 |  |  | 7 | 7 | 7 |  |  |

IHR: International Health Regulations (2005); NFP: National Focal Point.
<sup>a</sup> Facilitate communication and collaboration between NFPs and WHO International Food Safety Authorities Network (INFOSAN) emergency contact points during a foodborne illness emergency event.
<sup>b</sup> To examine protocols when working with non-health actors, particularly national disaster management agencies.
<sup>c</sup> Improve collaboration with other agencies.
<sup>d</sup> Not a listed objective but still tested.
<sup>e</sup> Videoconferencing was introduced to IHR Exercise Crystal for the first time.
1. Test the WHO guide on IHR Communications and Duty Officer system.
2. Validate IHR verification process between Member States and WHO for a public health emergency of international concern.
3. Improve the engagement of WHO Country Offices in facilitating communication between NFPs and WHO IHR Contact Point.
4. Practice and evaluate the NFPs' understanding and use of IHR principles and obligations.

The study used descriptive statistics to summarize and describe the performance of IHR NFPs and Contact Points in the Western Pacific Region during IHR Exercise Crystal for six key variables relating to NFP functions (Table 2). For each variable, the number and percentage of countries meeting the criteria were calculated for each year and the mean was calculated for all years.

**Table 2.**
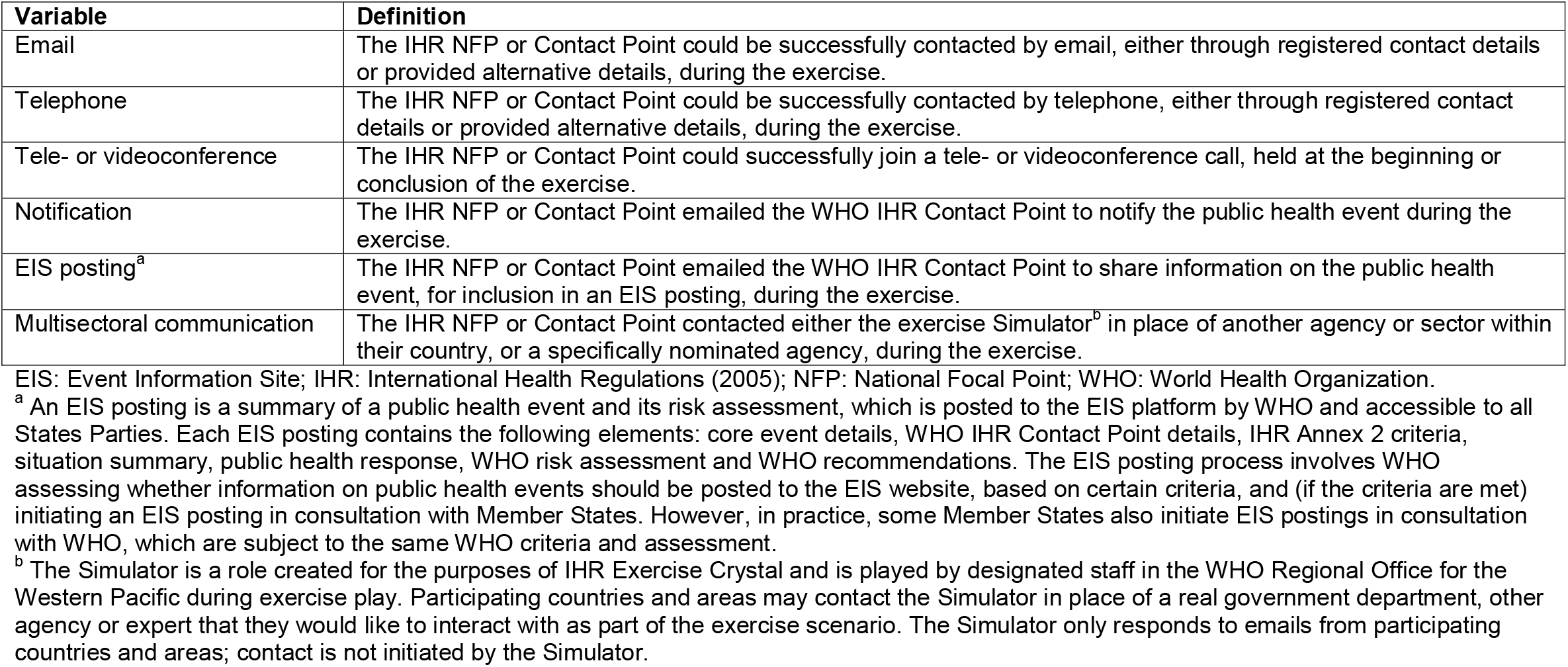
IHR Exercise Crystal variable definitions.

| Variable | Definition |
| --- | --- |
| Email | The IHR NFP or Contact Point could be successfully contacted by email, either through registered contact details or provided alternative details, during the exercise. |
| Telephone | The IHR NFP or Contact Point could be successfully contacted by telephone, either through registered contact details or provided alternative details, during the exercise. |
| Tele- or videoconference | The IHR NFP or Contact Point could successfully join a tele- or videoconference call, held at the beginning or conclusion of the exercise. |
| Notification | The IHR NFP or Contact Point emailed the WHO IHR Contact Point to notify the public health event during the exercise. |
| EIS posting<sup>a</sup> | The IHR NFP or Contact Point emailed the WHO IHR Contact Point to share information on the public health event, for inclusion in an EIS posting, during the exercise. |
| Multisectoral communication | The IHR NFP or Contact Point contacted either the exercise Simulator<sup>b</sup> in place of another agency or sector within their country, or a specifically nominated agency, during the exercise. |

EIS: Event Information Site; IHR: International Health Regulations (2005); NFP: National Focal Point; WHO: World Health Organization.
<sup>a</sup> An EIS posting is a summary of a public health event and its risk assessment, which is posted to the EIS platform by WHO and accessible to all States Parties. Each EIS posting contains the following elements: core event details, WHO IHR Contact Point details, IHR Annex 2 criteria, situation summary, public health response, WHO risk assessment and WHO recommendations. The EIS posting process involves WHO assessing whether information on public health events should be posted to the EIS website, based on certain criteria, and (if the criteria are met) initiating an EIS posting in consultation with Member States. However, in practice, some Member States also initiate EIS postings in consultation with WHO, which are subject to the same WHO criteria and assessment.
<sup>b</sup> The Simulator is a role created for the purposes of IHR Exercise Crystal and is played by designated staff in the WHO Regional Office for the Western Pacific during exercise play. Participating countries and areas may contact the Simulator in place of a real government department, other agency or expert that they would like to interact with as part of the exercise scenario. The Simulator only responds to emails from participating countries and areas; contact is not initiated by the Simulator.

## RESULTS

Between 2008 and 2024, IHR Exercise Crystal was held 16 times. No exercise was held in 2009; however, NFPs communicated frequently with the WHO IHR Contact Point on pandemic influenza A(H1N1), which was a public health emergency of international concern at the time. Additionally, a shared exercise was held with the Food and Agricultural Organization (FAO)/WHO International Food Safety Authorities Network (INFOSAN) in 2014. Twelve of the 16 exercise scenarios were related to outbreaks of respiratory viruses (Table 3). From 2008 to 2015, only States Parties (n = 27) to the IHR in the Western Pacific Region were invited to participate, with an average of 20 participating countries and areas (range 10–26) taking part each year. From 2016 onwards, all 37 countries and areas in the Region were invited to participate, with an average of 28 (range 14–35) taking part each year (Fig. 1). Mean participation for the period of 2008–2024 was 75% (Table 3).

**Table 3.**
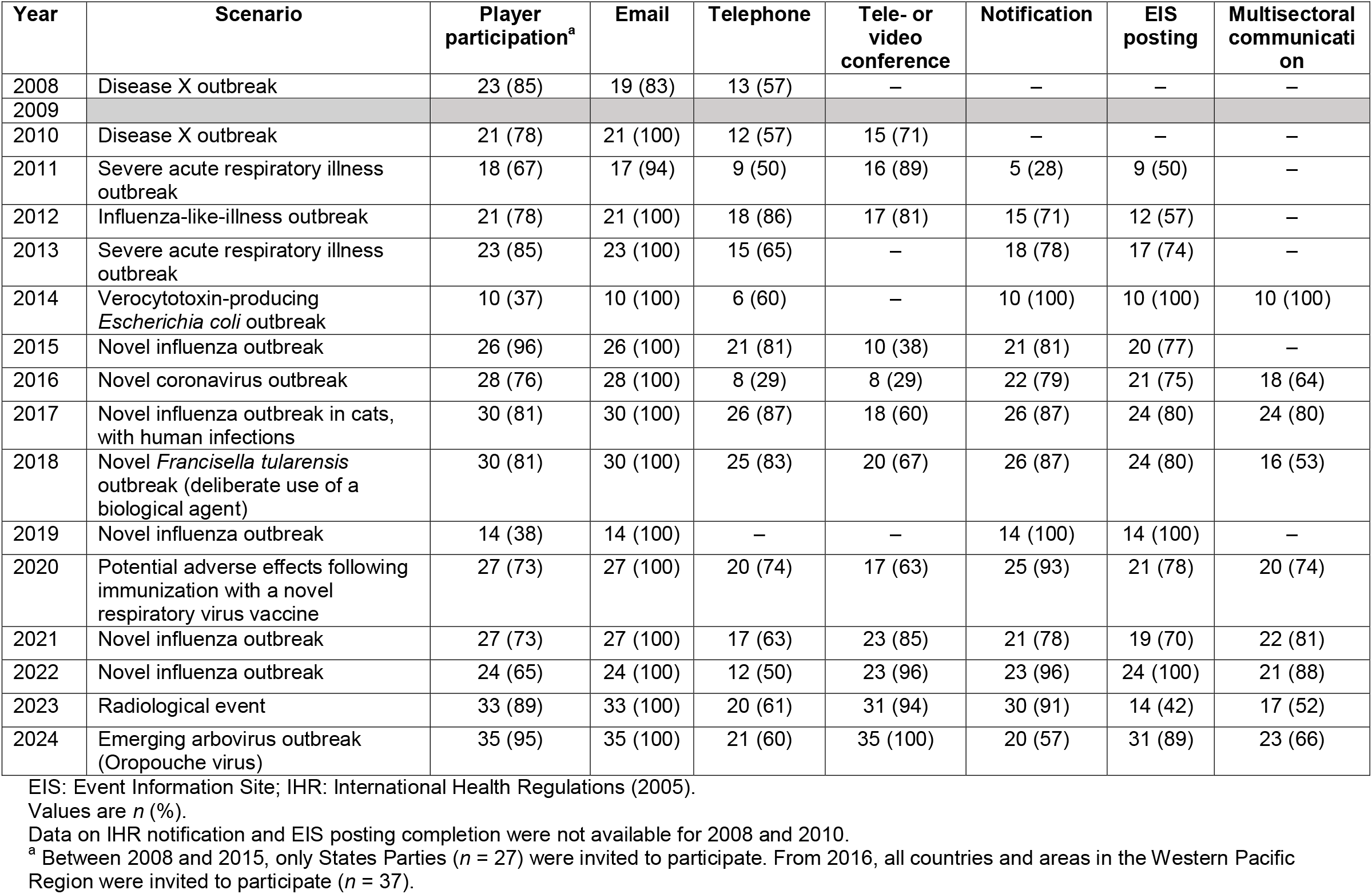
Summary of participating country and area performance during IHR Exercise Crystal by year, 2008–2024.

**Fig. 1.**
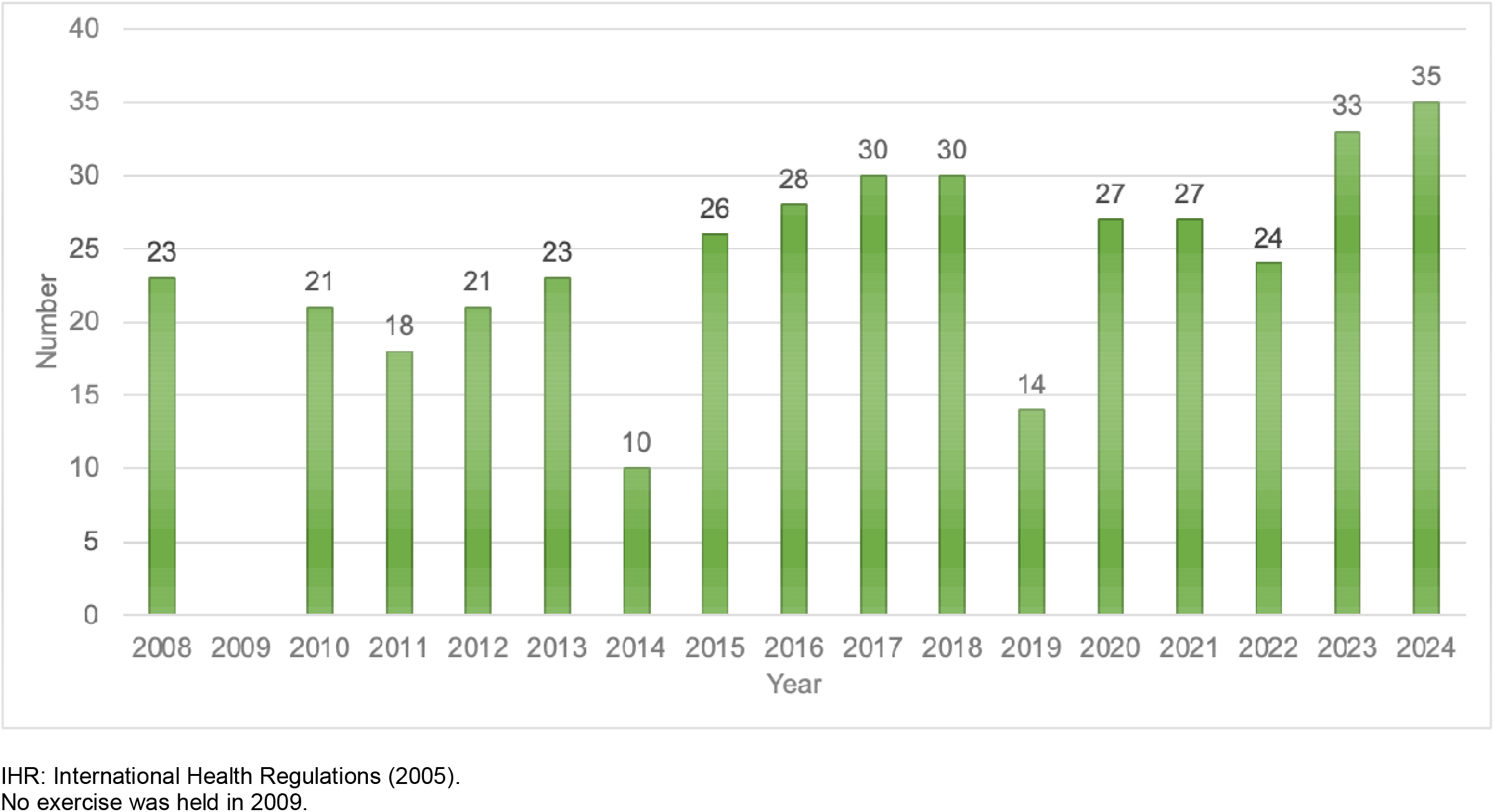
Number of countries and areas participating in IHR Exercise Crystal by year, 2008–2024

The proportion of participating countries and areas who were accessible via email was consistently high, with a mean of 99% for the period of 2008–2024 (Table 3). However, the proportion of participating countries and areas who were accessible via telephone has been suboptimal, with a mean of 64% for the period of 2008–2024 (Table 3). The proportion of participating countries and areas who joined a teleconference or a videoconference (first introduced in 2015) with the Regional Office improved over time, with 85% or more successfully attending between 2021 and 2024, for a mean of 73% over the entire period of 2008–2024 (Table 3).

The proportion of participating countries and areas notifying the simulated event to the WHO IHR Contact Point has improved over time, with some variation between years. Between 2008 and 2024, a mean of 80% of participating countries and areas notified the simulated event to WHO (Fig. 2). The proportion of participating countries and areas who have contributed information to an Event Information Site (EIS) posting has typically followed a similar trend to that of notifications, with some variation in 2023 and 2024. Between 2008 and 2024, a mean of 77% of participating countries and areas contributed information to an EIS posting (Fig. 2).

**Fig. 2.**
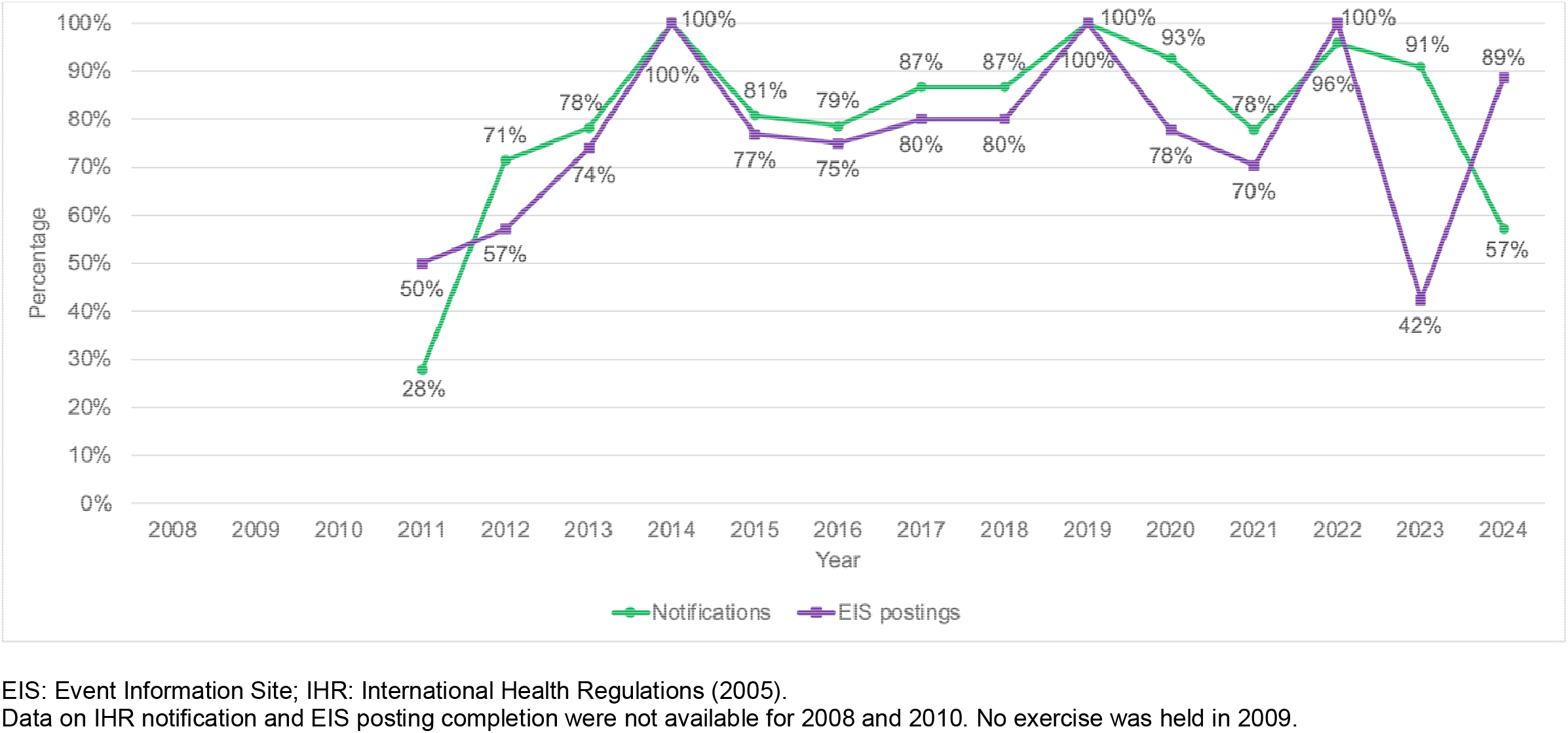
Percentage of participating countries and areas completing IHR notifications and EIS postings during IHR Exercise Crystal by year, 2008–2024

Data for the variable on multisectoral communication was limited for the period of 2008–2024, as this was not a listed objective of IHR Exercise Crystal for six of the 16 years (Table 1). Though no clear trend in NFP capacity to conduct multisectoral communication was able to be observed over time, the mean performance was 73% for the nine years of available data (Table 3). Furthermore, where data were available, multisectoral communication scored the lowest for the radiological, vector-borne disease and non-influenza respiratory disease outbreaks caused by *Francisella tularensis* and novel coronavirus, and highest for infectious disease outbreaks involving a novel influenza or gastroenteritis (Table 3).

## DISCUSSION

Since its beginning in 2008, IHR Exercise Crystal has seen strong participation from Member States in the Western Pacific Region, and several key NFP performance indicators have been tracked over time. Our analysis of these indicators demonstrates that the accessibility of NFPs using registered contact details has continued to be a challenge. Email has proven to be the most effective means of communication, with a mean of 99% of NFPs successfully being contacted by email, compared to a mean of 64% of NFPs successfully being contacted by telephone. The proportion of NFPs who are able to join the exercise by tele- or videoconferences has improved over time. These trends are consistent with previous findings on NFP accessibility in the Region^12^ and demonstrate that while NFP performance has continued to improve, there remains a need to regularly (at least annually) test and update NFP contact details. This is supported by a global survey of States Parties conducted in 2019, which found that communications were one of four critical areas where NFPs experienced challenges.^13^

Similarly, our finding that the proportion of NFPs notifying simulated public health events to the WHO IHR Contact Point and the proportion of NFPs contributing information to EIS postings have improved over time is consistent with findings from other studies.^12,13,15^ In one study, most (96%) NFPs reported that they are familiar with how to contact their designated WHO IHR Contact Point and that they have the necessary content expertise to discuss a notifiable event with the WHO IHR Contact Point.^13^ In another study, 88% (*n* = 112) of NFPs reported that they had excellent or good knowledge of the Annex 2 decision-making instrument, and either excellent (30, 22.9%) or good (58, 44.2%) ability to assess potential public health emergencies of international concern under Annex 2.^14^ The strong performance in this NFP capacity is likely due to the long-term investments made by WHO and States Parties in institutionalizing the use of Annex 2 through training, guidance documents or standard operating procedures (SOPs) and the development of legal, regulatory or administrative provisions supporting its use.^15–18^

Our analysis did not find a clear trend in NFPs’ capacity to conduct multisectoral communication; however, on average 73% of NFPs communicated with another sector or agency during exercise play. It was previously identified that NFPs experience challenges in intersectoral collaboration within their countries, including limited access to or a lack of cooperation from key relevant ministries,^13^ and that NFPs were not sufficiently empowered to carry out their functions,^13,19,20^ which creates difficulties in engaging directly with other agencies or sectors and in triggering decision-making processes by national health authorities.^20^ For this reason, it has been recommended that States Parties establish a National IHR Authority that will focus on the implementation of the IHR across sectors, recognizing that the core capacities required extend beyond the health sector.^20,21^ Though the role and functions of NFPs do not change under the June 2024 IHR amendments,^3^ the added requirement to designate or establish a National IHR Authority means that it will be even more important to ensure the clear delineation of roles and responsibilities and for States Parties to develop, test and maintain up-to-date SOPs, particularly in relation to multisectoral communication and coordination.

States Parties should also continue strengthening and empowering NFPs to conduct their core functions of IHR communications. Tools such as SPAR and IHR Exercise Crystal can guide continuous improvement of NFP functionality.^22^ Furthermore, APHSAF advocates for strengthening the mandate and capacities of NFPs, ensuring that NFP capacities are prepared and ready to respond to public health emergencies (through regular testing, for example), and enhancing communication, information sharing and coordination between the National IHR Focal Point system and emergency contact points for other areas and sectors, as well as between countries.^4^

Limitations of this analysis include the inability to capture multisectoral communications that occur outside of observed exercise email communications and variation in IHR Exercise Crystal monitoring and evaluation methods over time. Triangulation with multiple data sources, such as SPAR, is therefore important to consider when analysing and interpreting the results of simulation exercises.

## Conclusion

Between 2008 and 2024, States Parties in the Western Pacific Region demonstrated improved NFP capacities in the areas of IHR notification, contributing information to EIS postings, and tele- and videoconferencing. Continued strengthening is required, particularly in the areas of NFP accessibility and multisectoral communications. Simulation exercises like IHR Exercise Crystal are one tool that States Parties can use to assess NFP capacities and guide improvements. NFP functions do not change in the context of the 2024 IHR amendments and the designation or establishment of a National IHR Authority; however, States Parties should clearly define responsibilities and develop and test operational procedures to ensure that NFPs continue to function without disruption. This is critical to advancing health security and IHR implementation in the Region.

## Data Availability

All data produced in the present study are available upon reasonable request to the authors

## ACKNOWLEDGEMENTS

The authors would like to thank colleagues from Member States in the Western Pacific Region, WHO country offices and WHO headquarters for their participation and support in IHR Exercise Crystal.

## ETHICS STATEMENT

This regional analysis consists of a review and synthesis of publicly available public health data. It does not involve human participants, identifiable personal data, or interventions. Based on organizational ethical review policies, such activities do not require formal ethics approval.

